## Supplementary Tables and Figures for "Genome sequencing and transcriptome profiling in twins discordant for Mayer-Rokitansky-Küster-Hauser syndrome"

**Supplementary Table 1. Multi-sample analysis**

| Twin Pair | Total variant number | Multi-sample dominant filter | Quality filter* | Modified** multi-sample dominant filter + target region filter | Modified multi-sample dominant filter + target region filter quality filter | After literature review and manual variant evaluation |
|---|---|---|---|---|---|---|
| Pair 1 | 387411 | 2 | 0 | 20 | 10 | 0 |
| Pair 2 | 384229 | 4 | 1 | 32 | 5 | 1 |
| Pair 3*** | 367383 | 12 | 2 | 21 | 3 | 0 |
| Pair 4 | 380390 | 9 | 0 | 22 | 2 | 0 |
| Pair 5 | 388670 | 3 | 1 | 19 | 2 | 0 |

This table shows the reduction of variants found in GSVar due to the implementation of different filter criteria in a multi-sample analysis comparing tissue and blood samples of the affected twin to blood samples of the healthy twin.

*qual: 250 | depth: 0 | mapq: 55 | strand bias: 20 | allele balance: 80
**without high, moderate, low impact filter
*** only blood of MRKH-twin and healthy twin were available for analysis

**Supplementary Table 2. Blood-Tissue analysis**

| Twin Pair | Total variant number | Modified multi-sample dominant filter** + target region filter | Modified multi-sample dominant filter + target region filter + quality filter* | After manual variant evaluation |
|---|---|---|---|---|
| Pair 1 | 359563 | 46 | 9 | 0 |
| Pair 2 | 359162 | 60 | 9 | 0 |
| Pair 4 | 354972 | 37 | 6 | 0 |
| Pair 5 | 362612 | 45 | 11 | 0 |

This table depicts the reduction of variants found in GSVar due to the implementation of different filter criteria in an analysis comparing tissue and blood of affected twin.

*qual: 250 | depth: 0 | mapq: 55 | strand bias: 20 | allele balance: 80
**without high, moderate, low impact filter

### Single sample analysis

**Supplementary Table 3. Single-sample analysis**

| Twin Pair | Total variant number | Allele-frequency based filter | Target Region Filter | After literature review and manual variant evaluation |
|---|---|---|---|---|
| Pair 1 | 354179 | 455 | 28 | 1 |
| Pair 2 | 367131 | 515 | 36 | 1 |
| Pair 3 | 385106 | 503 | 26 | 1 |
| Pair 4 | 344744 | 427 | 18 | 0 |
| Pair 5 | 374078 | 475 | 27 | 0 |

This table shows the reduction of variants found in GSVar after implementing different filter criteria in a single-sample analysis of tissue from the affected twin.

#### Supplementary Figure 1

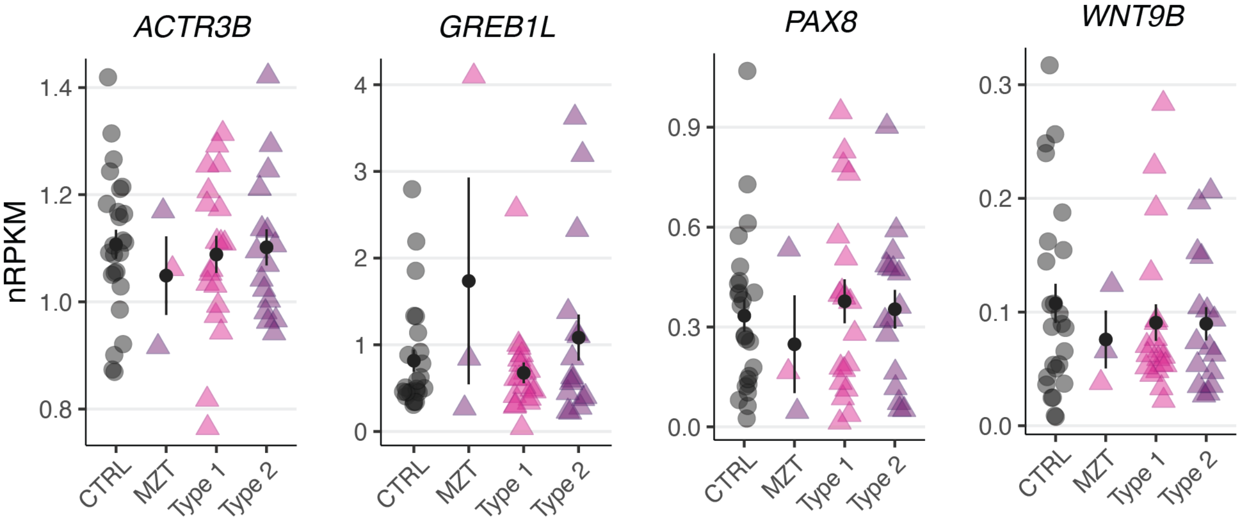

#### Supplementary Figure 2

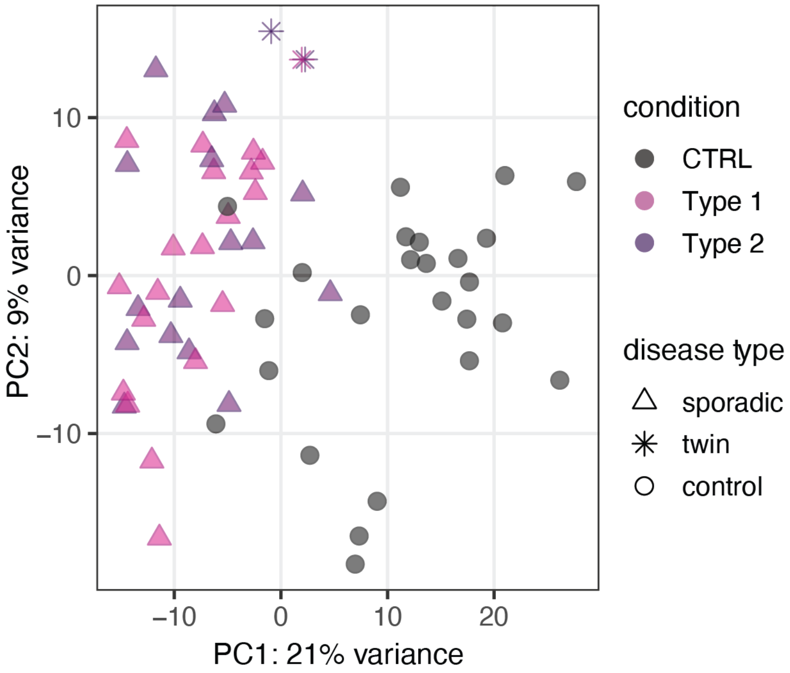

### Supplementary Figure 3

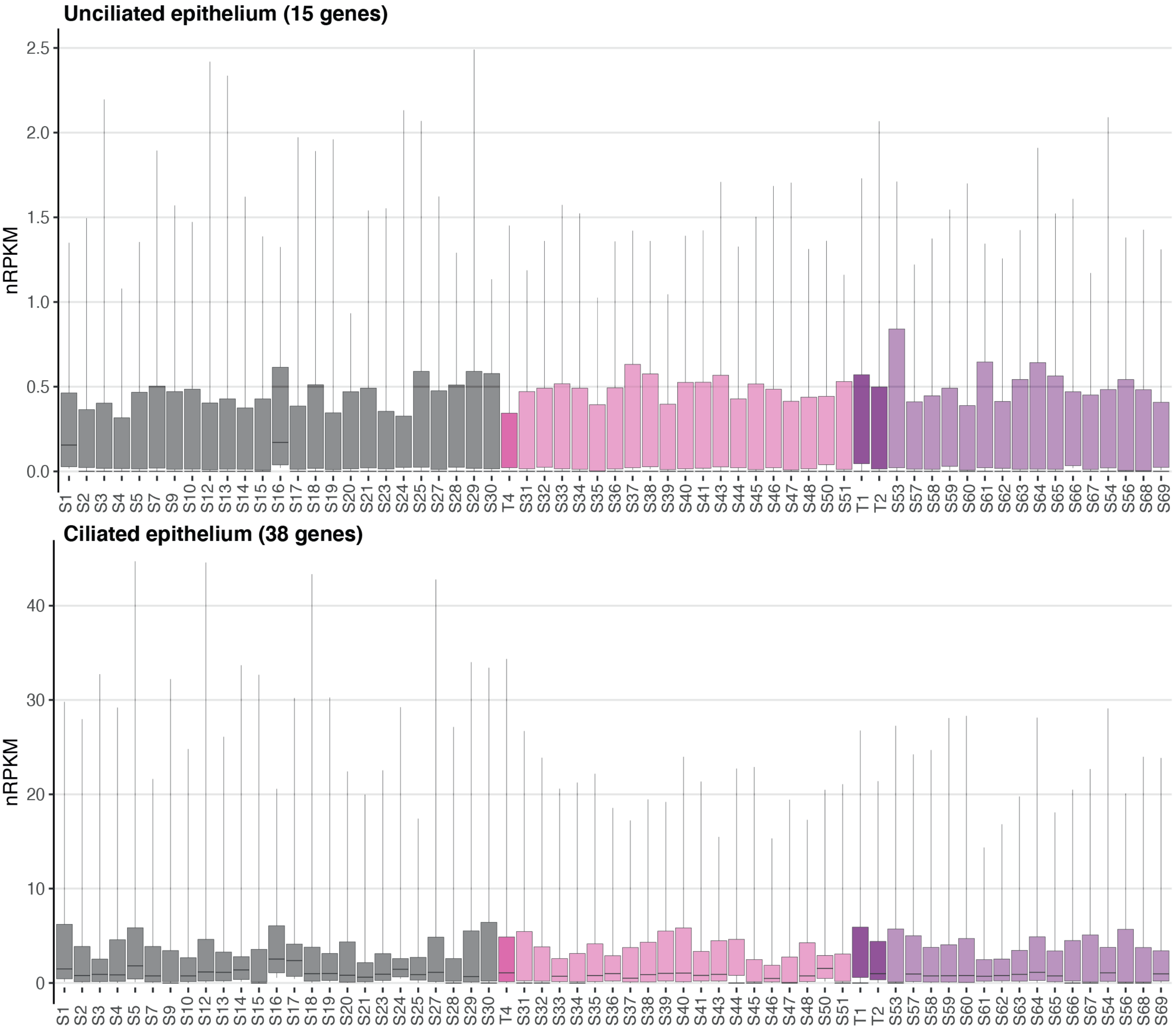
